# Use of machine learning to predict hypertension-related complication outcomes of varying severity

**DOI:** 10.1101/2020.10.30.20169615

**Authors:** Jasmine M. McCammon, Sricharan Bandhakavi, Doreen Salek, Zhipeng Liu, Xianglian Ni, Natalie Benner, Ronda Rogers, Hollie Yoder, Shelley Riser, Farbod Rahmanian

## Abstract

**Objective:** A challenge in hypertension-related risk management is identifying which people are likely to develop future complications. To address this, we present administrative-claims based predictive models for hypertension-related complications.

**Materials and Methods:** We used a national database to select 1,767,559 people with hypertension and extracted 112 features from past claims data based on their ability to predict hypertension complications in the next year. Complications affecting kidney, brain, and heart were grouped by clinical severity into three stages. Extreme gradient boosting binary classifiers for each stage were trained and tuned on 75% of the data, and performance on predicting outcomes for the remaining data and an independent dataset was evaluated.

**Results:** In the cohort under study, 6%, 17%, and 7% of people experienced a hypertension-related complication of stage 1, stage 2, or stage 3 severity, respectively. On an independent dataset, models for all three stages performed competitively with other published algorithms by the most commonly reported metric, area under the receiver operating characteristic curve, which ranged from 0.82-0.89. Features that were important across all models for predictions included total medical cost, cost related to hypertension, age, and number of outpatient visits.

**Discussion:** The model for stage 1 complications, such as left ventricular hypertrophy and retinopathy, is in contrast to other offerings in the field, which focus on more serious issues such as heart failure and stroke, and affords unique opportunities to intervene during earlier stages.

**Conclusion:** Predictive analytics for hypertension outcomes can be leveraged to help mitigate the immense healthcare burden of uncontrolled hypertension.

**LAY SUMMARY:** As the leading preventable risk factor for morbidity and mortality in the world, identifying which people with hypertension are likely to exacerbate is critically important for development of effective intervention strategies. Here we present a suite of predictive models that can predict future risk of development of hypertension-related complications. To have utility for triaging as well as identifying mild cases before they progress to critical end phases, the models predict three different stages of severity of hypertension-related complications. Our algorithms utilize variables calculated for the most recent 12 months, and predict probability of a hypertension-related complication for the next 12 months using administrative claims as the data source. Because the types of complications that have been analyzed can also result from comorbidities besides hypertension, such as diabetes and hyperlipidemia, these diagnoses are included as variables. Other variables pertain to demographic characteristics, prescription information, relevant procedures, and utilization patterns. Overall, all three models exhibited strong predictive performance. The ability to use straightforward variables found in claims data to predict future risk of disease-related complications, complemented with targeted clinical intervention strategies, has the potential to reduce cost of care and improve health outcomes for the many people living with hypertension.

## BACKGROUND AND SIGNIFICANCE

Hypertension (HTN) is the chronic elevation of blood pressure due to changes in cardiac output and/or systemic vascular resistance, which can lead to organ damage and mortality if left unmanaged. The overall prevalence of HTN is high, nearly a third of the global adult population, making HTN one of the basic cost drivers in healthcare systems [1]. Despite continuing advances in treatment options, HTN remains uncontrolled in more than half of the population on medication [2]. As a result of its high prevalence and difficulty to adequately manage, HTN is the leading preventable risk factor globally for premature death and disability [3].

The contribution of HTN to morbidity and mortality is due to the long term effects of chronic, unmanaged high blood pressure. HTN itself is without symptoms, but left uncontrolled, will lead to systemic organ damage over time [4]. Initial stages of damage such as microalbuminuria and left ventricular hypertrophy are subtle and may go unrecognized until they’ve progressed to more serious complications such as chronic kidney disease and heart failure [5]. Estimates suggest that preventing hypertension could reduce cardiovascular death by 38% in females and 30% among males [6]. Another study found that 48% of population risk for stroke could be attributed to HTN, the highest of all factors examined [7]. After diabetes, HTN is the second leading cause of end-stage renal disease [8]. Therefore, the ability to predict who with HTN is likely to exacerbate, or present with HTN-related complications, before such events occur is a powerful opportunity to mitigate some of this morbidity and mortality burden.

We found a number of studies that used machine learning algorithms to predict various complications associated with HTN. Diverse data sources, including EHR, clinical trial data, and national healthcare databases from the US, Korea, China, and Europe were utilized to predict complications such as cardiovascular disease, dementia, myocardial infarction, acute coronary syndrome, cerebrovascular disease, stroke, kidney disease, heart failure, coronary heart disease, ischemic heart disease, and cardiovascular death [9-17]. In general, the number of people included in these cohorts ranged from 3,395 to 133,176; only three included people below the age of 40 and one did not report age ranges [9, 10, 16, 17]. The area under the receiver operating characteristic (AUROC) was the most commonly reported evaluation metric, ranging from 0.56-0.84, with an average of 0.71 across seven studies [9-13, 15, 17].

From this, we saw an opportunity to contribute to this strong predictive modeling work by exploiting a larger dataset, a national collection of administrative claims from commercially-insured individuals, with hypertensive members in the millions. We refer to this source as the CCAE (Commercial Claims and Encounters) database within this manuscript. Including people in the age range of 18-64 represents a lower risk population for complications, but more opportunities for targeted intervention before the risks may become unmanageable. Our independent test set did include people over 65 to assess the model’s generalizing ability to this demographic. In addition, rather than focusing on a specific type of complications, a specific organ system, or a composite of severe complication outcomes as has been done in previous studies, we chose to build separate models to predict the complications that reflect the progression of chronic HTN. Composite outcomes across organ systems were grouped into three stages of increasing clinical severity as described by Messerli and colleagues [18]. Interventions during earlier signs and symptoms of HTN complications are more likely to be successful than interventions at later stages and may help to prevent later stage HTN complications as well [19]. Features thought to correlate directly or indirectly with HTN-related complications were calculated for the intake window, a 12-month period before the year for prediction, and extreme gradient boosting (XGBoost) classifiers were trained to predict the likelihood of these outcomes in the following year. Overall, model performance was strong on the independent dataset (AUROC = 0.82-0.89), comparable or better than the previously published algorithms mentioned above, in addition to having the benefits of providing early intervention opportunities. These models exhibit practical utility from a care management perspective to triage and manage hypertensive populations, which are inherently large, for different levels of HTN complication severity.

## MATERIALS AND METHODS

### Study design and sample data

We used administrative claims data generated between January 1, 2016 to December 31, 2017 from commercially-insured individuals in the CCAE database. The cohort of study was selected based on data from 2016 and HTN-related complication events were identified in 2017 data. To be included in the cohort, members were presumed to have HTN based on their medical and pharmacy claims: they either had to have at least two claims 14 or more days apart with a HTN diagnosis or at least one claim with a HTN diagnosis and at least two pharmacy claims with anti-HTN medications (for medical codes used see Supplementary Table S1). They also had to be continuously enrolled in 2016 and 2017 in medical and pharmacy benefits and be at least 18 years old. We also explored cohorts that did and did not include members that had complication events during the intake window of 2016.

Features for each cohort member were then calculated for 2016 to use for predicting hypertension complications in 2017. An independent dataset of administrative claims from an independent payer not in the CCAE database was used to process data in the exact same manner and validate final model performance.

### Outcome variable: Defining a hypertension complication

Outcomes were identified during the prediction window of January 1, 2017 to December 31, 2017. In order to maximize applications and utility, we chose to build three different models predicting different levels of complication severity across different target organ systems (Figure 1). Stage 1 complications are the least severe, and include proteinuria, left-ventricular hypertrophy, and retinopathy. Stage 2 complications, the next step in progression of unmanaged hypertension, include chronic kidney disease, coronary artery disease, and transient ischemic attack. The most severe complications, stage 3, include end-stage renal disease, heart failure, and stroke. As a result, stage 1 and stage 2 models can be used to target people in the early stages of HTN progression, while predictions for stage 3 complications can be used to triage for likelihood of severe HTN exacerbations for those that have already progressed to an advanced disease state. ICD-10-CM diagnosis codes [20] used to identify these complication events can be found in Supplementary Table S2. These labels were treated as non-mutually exclusive when training each model. In other words, the same cohort was used to fit all three models and members were not excluded if they had multiple stages of complications during the prediction window.

**Figure 1.**
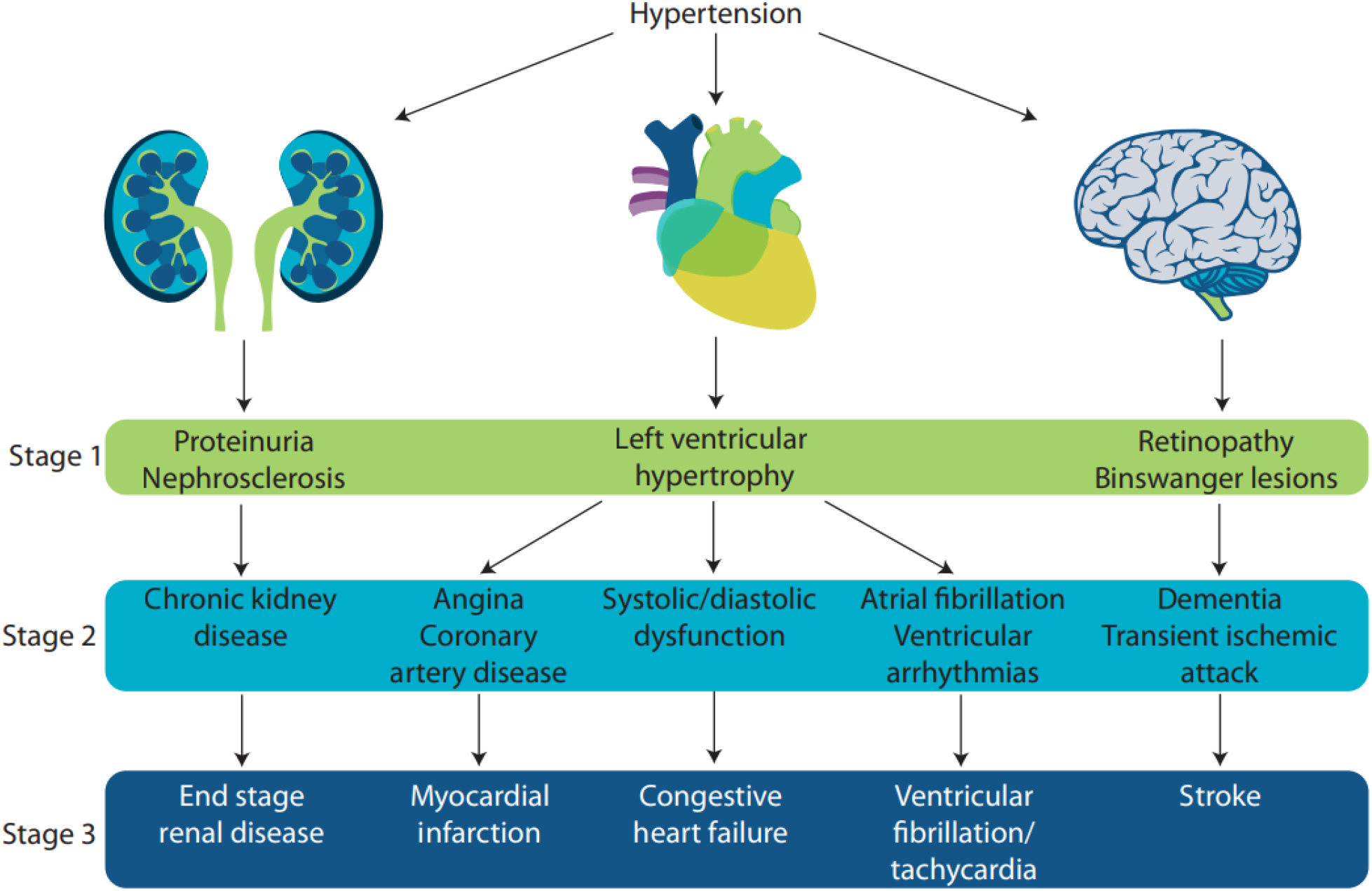
Hypertension complications. Target organs affected are kidney, heart, and brain. Complications progress with increasing severity and can ultimately result in mortality. Adapted from [18].

### Feature candidates

We compiled 112 different features based on previous literature and clinical information, all during the intake window of January 1, 2016 to December 31, 2016. To ascertain the feature of whether each member was taking anti-hypertensives for the first time in 2016, what we call a “progressor” [21], pharmacy data from 2015 was also used, but we allowed for progressor status as unknown if they were not enrolled in 2015. Demographic features were age and gender. Diagnoses included comorbidities common with hypertension and known to contribute to increased likelihood of disease exacerbation, such as diabetes and hyperlipidemia, as well as other conditions that could contribute to the complication set. For example, heart valve disorders could also lead to arrhythmias. The Charlson Comorbidity Index was calculated as well [22]. Prescription features pertained to anti-hypertensive drug classes: diuretics, calcium channel blockers, agents acting on the renin-angiotensin system, beta blockers, vasodilators, and adrenergic antagonists. Mono- and polytherapies as well as medication changes were evaluated [21]. Medications indicative of comorbidities, such as metformin for diabetes, were included, as well as medications known to interfere with anti-hypertensives, like steroids [23]. In the absence of clinical data, counts of unique dates with particular procedure codes were used in place of laboratory results, including tests for HbA1c and creatinine levels. Finally, utilization was assessed as counts of unique visit dates in different care settings: outpatient (OP), inpatient (IP), and emergency department (ED), as well as two cost features: total medical spending for the year and spending for claims with hypertension diagnoses.

### Machine learning approaches and prediction performance evaluation

Several algorithm architectures were tested, and we decided to use the XGBoost classifier [24], which had the best performance during initial data exploration phases. Three separate models were constructed for each stage to allow flexibility in fine-tuning for the most accurate predictions of each stage of complication. For model training and evaluation, data were split 75%-25% for training and test sets using stratified sampling. Model hyperparameters tuned with random grid search and cross validation were learning rate, maximum depth, minimum child weight, and subsample rate.

Model performance was evaluated with several metrics, including area under the receiver operating characteristic (AUROC), area under the precision-recall curve (AUPRC) [25], sensitivity, specificity, negative predictive value (NPV), positive predictive (PPV) value, F1 score, and number needed to evaluate (NNE) – or the number of people needed to evaluate to get one person with the true outcome – on the test set and an independent dataset. The optimal threshold for prediction cutoff was chosen to balance sensitivity and specificity as identified by the Youden index [26]. We also report feature importance for the top 20 most important predictors for each model.

Analyses and visualizations were done using Python, version 2.7, with the scikit-learn, numpy, pandas, and seaborn libraries (Python Software Foundation).

## RESULTS

### Descriptive statistics of the case population

There were 1,767,559 members in the cohort of study; of these, 396,607 (22.4%) had at least one HTN-related complication of any stage. Stage 2 complications were most common with 303,150 members (17.1% of cohort), while stage 1 (99,829) and stage 3 (131,126) were less so (5.6% and 7.4%, respectively). Some members experienced complications from two or all three stages (6.5%) during the prediction window. There was a lower percentage of females (41.3% vs. 49.0%) as well as a higher average age (54.7 vs. 51.6 years old) among those that experienced a HTN-related complication versus those that did not (Figure 2). Because more than half the people who experienced a stage 1 complication also experienced a stage 2 and/or stage 3 complication, and more than a quarter of people who had a stage 2 complication also had a stage 3 complication, the summary statistics for the features below pertain to mutually exclusive roll-up categories where a person was counted for his or her most severe complication stage.

**Figure 2.**
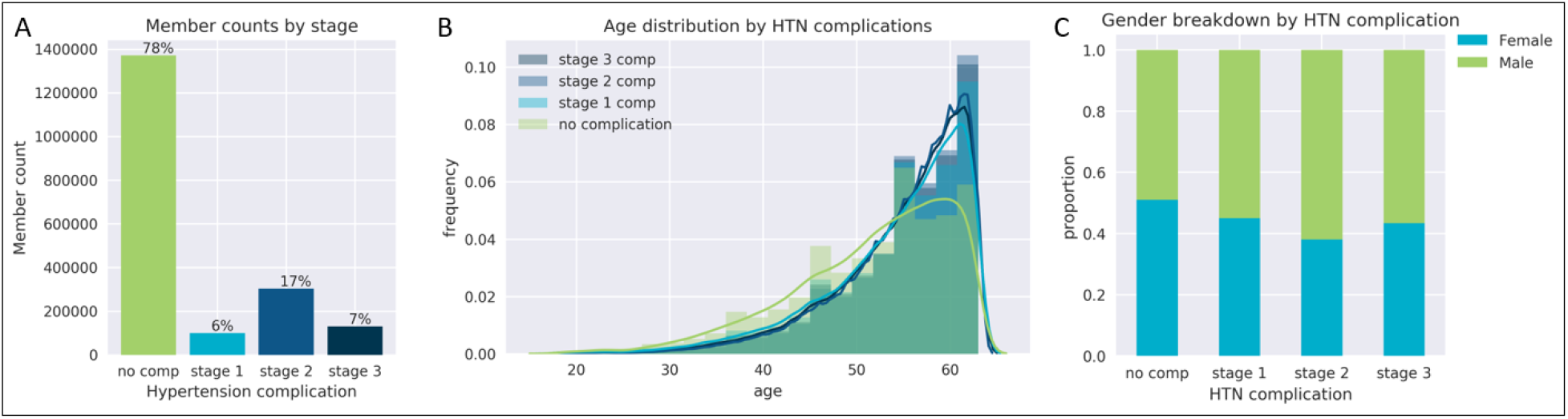
Hypertension complication prevalence and demographic summary. (A) Member counts by complication stage during prediction window. Members can experience complications from multiple stages, but are only counted as ‘no complication’ status if they do not experience complications of any stage. (B) Distribution of ages by complication status shows people that do not experience a complication tend to be slightly younger. This study design did not include people below the age of 18, and because it utilized administrate data from commercially insured individuals, does not include anyone over the age of 65. (C) Proportion of males and females by complication status shows roughly equal percentages of both sexes in the no complication group, but higher proportions of males in all three complication stage groups. Comp, complications; HTN, hypertension.

Select diagnosis and prescription features as they relate to HTN complication stages are shown in Figure 3. These were chosen as they are among the top 20 most important features for one or more of the models. Summary statistics of all diagnosis- and prescription-related features are in Tables 1 and 2, respectively. Charlson Comorbidity Index [22], a discrete risk score, increased as complication severity increased (Figure 3B). For diagnosis-related features scored categorically by the presence of at least one code (n=60), all but anxiety disorder and hemochromatosis were proportionally higher in all three of the complication stage groups compared to those that did not have any complications. Thirty-one of these diagnoses increased proportionally with increasing complication severity (Table 1). Discrete prescription features, such as number of different anti-hypertensive classes and number of anti-hypertensive treatment adjustment rounds, positively correlated with complication severity (Figure 3C). For prescription features scored categorically by the presence of at least one code (n=35), 20 were proportionally higher in all complication stage groups compared to those without any hypertension-related complications. Six prescription predictors decreased with complication severity (Table 2).

**Table 1.**
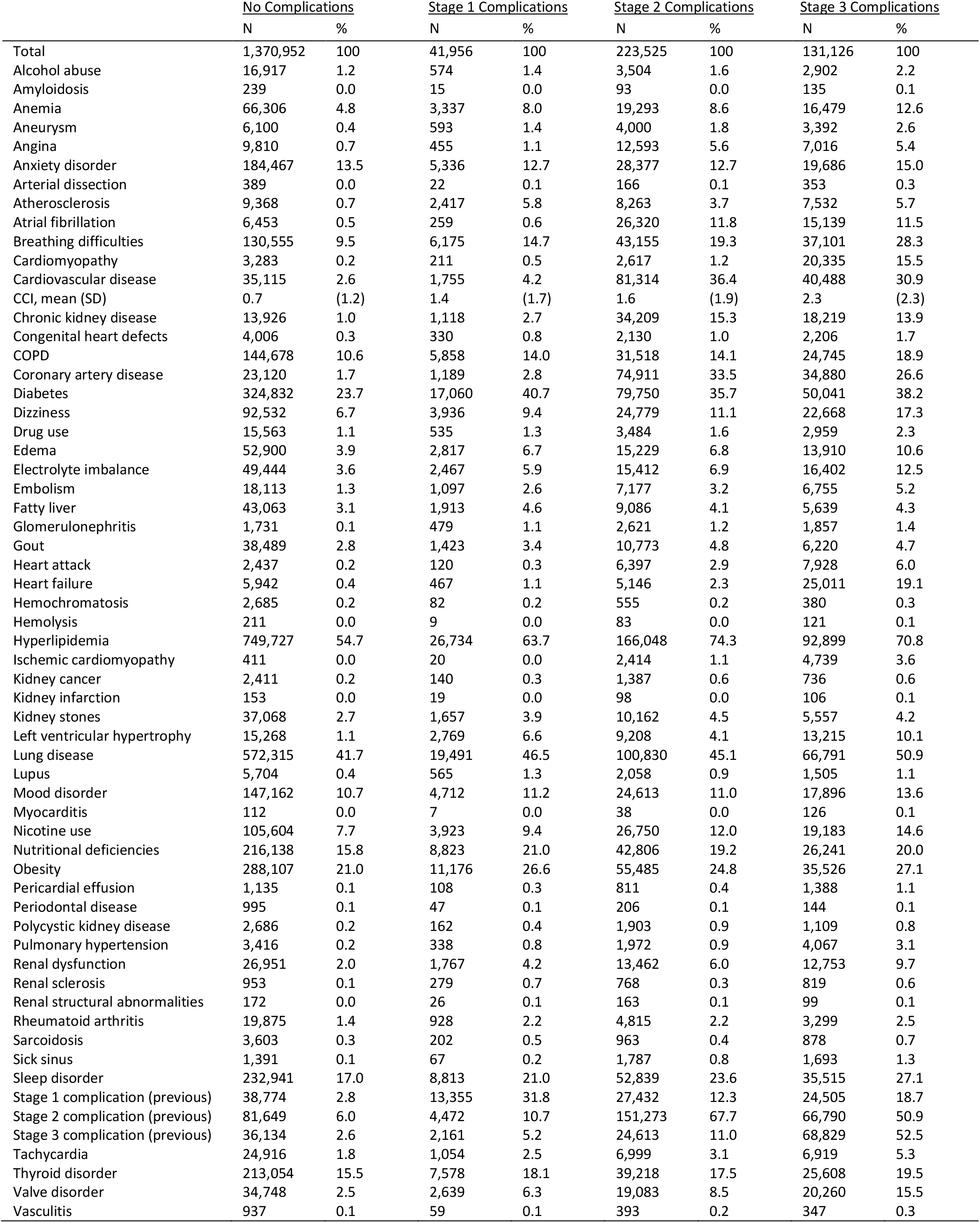
Diagnosis-related features for four groups: those that experienced no hypertension-related complications, stage 1, stage 2, or stage 3 complications. Member counts and proportions for people in each group are shown for most features, scored as presence or absence of at least one code, unless the feature is discrete, in which case mean and standard deviation are presented. CCI, Charlson Comorbidity Index; COPD, chronic obstructive pulmonary disease; SD, standard deviation.

**Table 2.**
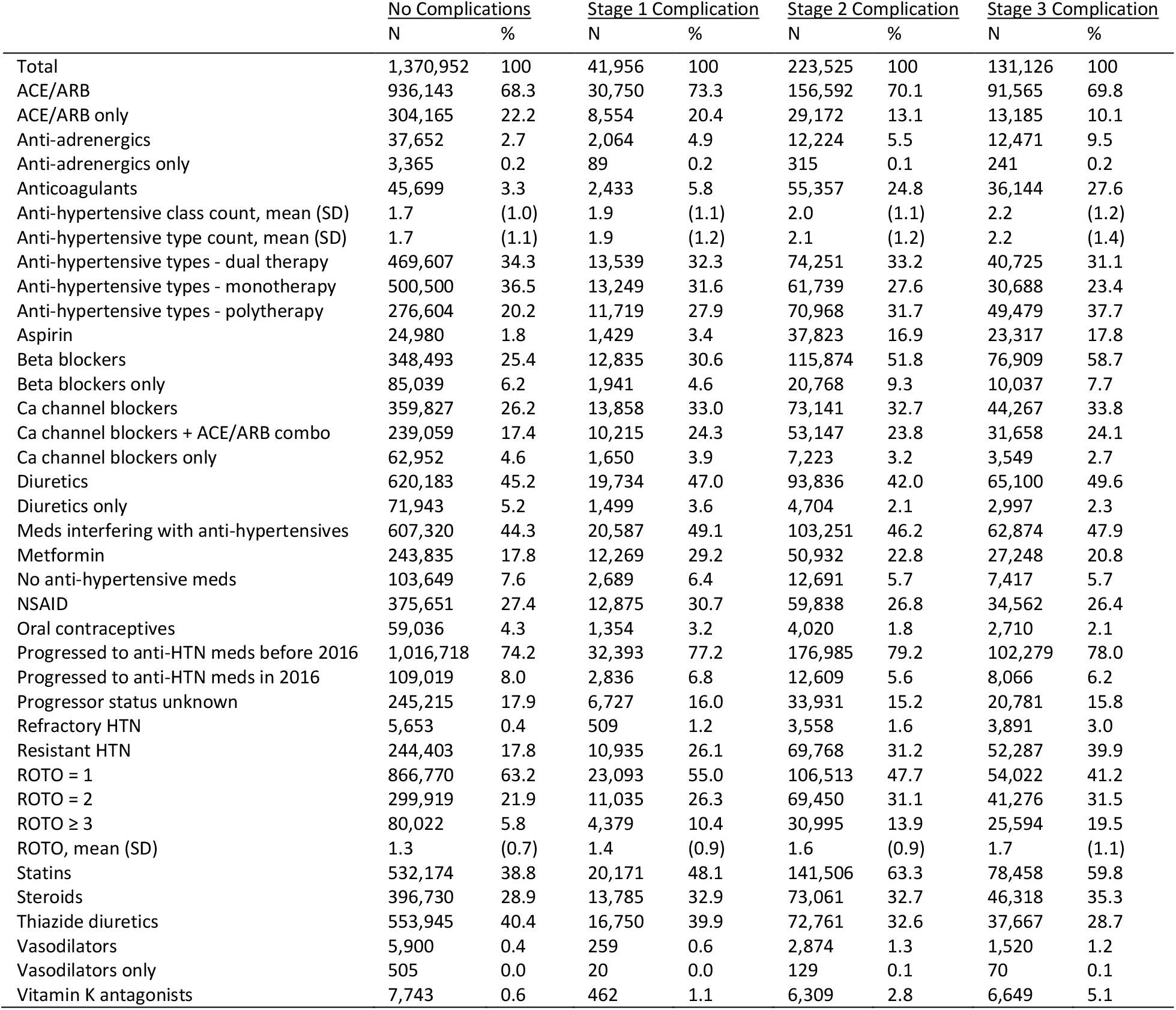
Predictors pertaining to pharmacy claim data. An anti-hypertensive “type” refers to its 5 digit ATC4 hierarchy code, whereas “class” refers more broadly to beta blockers, diuretics, calcium channel blockers, etc., or its 3 digit ATC4 hierarchy code. Predictors are scored categorically and presented as proportions of total number of members in that complication class, unless denoted with “mean (SD),” in which case they are discrete features and summary statistics of mean and SD are reported. Abbreviations: ACE, angiotensin converting enzyme inhibitor; ARB, angiotensin II receptor blocker; Ca, calcium; HTN, hypertension; NSAID, nonsteroidal anti-inflammatory drug; ROTO, rounds of treatment options; SD, standard deviation.

**Figure 3.**
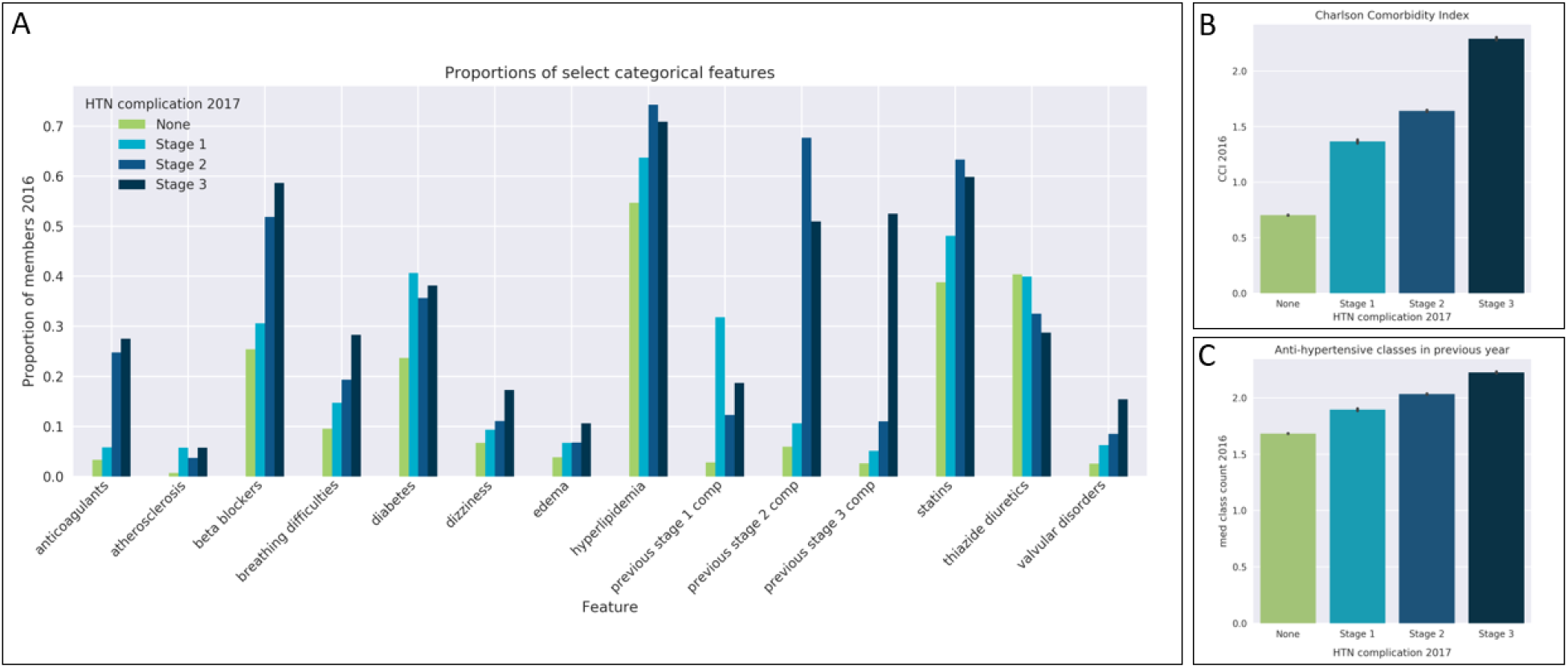
Selected diagnosis and medication features. (A) Features during intake year (2016) shown as proportion of members in each group with the presence of at least one diagnosis or drug code belonging to that category. A person is grouped according to his or her most severe complication stage experienced in 2017. (B) The Charlson Comorbidity Index, a composite weighted risk score of 19 different chronic diseases, increases with increasing complication severity. (C) The count of different anti-hypertensive drug classes (diuretics, calcium channel blockers, agents acting on the renin-angiotensin system, beta blockers, vasodilators, and adrenergic antagonists) increases with increasing complication severity. Bars represent 95% confidence intervals.

Counts of relevant procedure codes (n=6 predictors) and utilization visits and costs (n=5 predictors) were all discrete features, and means and SDs of these predictors by complication stage can be found in Table 3. The mean for all of these features was lowest in the complication group versus all other complication stages. Mean utilization across all categories positively correlated with complication stage severity, as did all procedure code counts except for HbA1c and at home blood pressure monitor (Table 3, Figure 4).

**Table 3.**
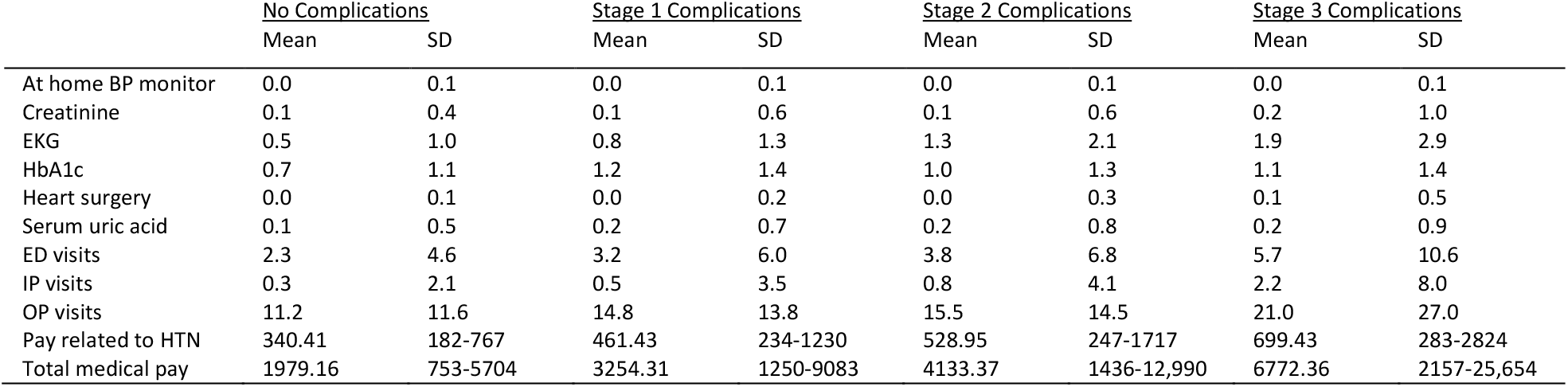
Features related to utilization and procedure counts. For each complication group, mean and standard deviation are reported, except for pay features, where median and interquartile range are reported. For the procedures and visit types listed, unique claim dates were counted per person during the intake window. BP, blood pressure; EKG, electrocardiogram; HbA1c, hemoglobin A1c; ED, emergency department; IP, inpatient; OP, outpatient; HTN, hypertension.

**Figure 4.**
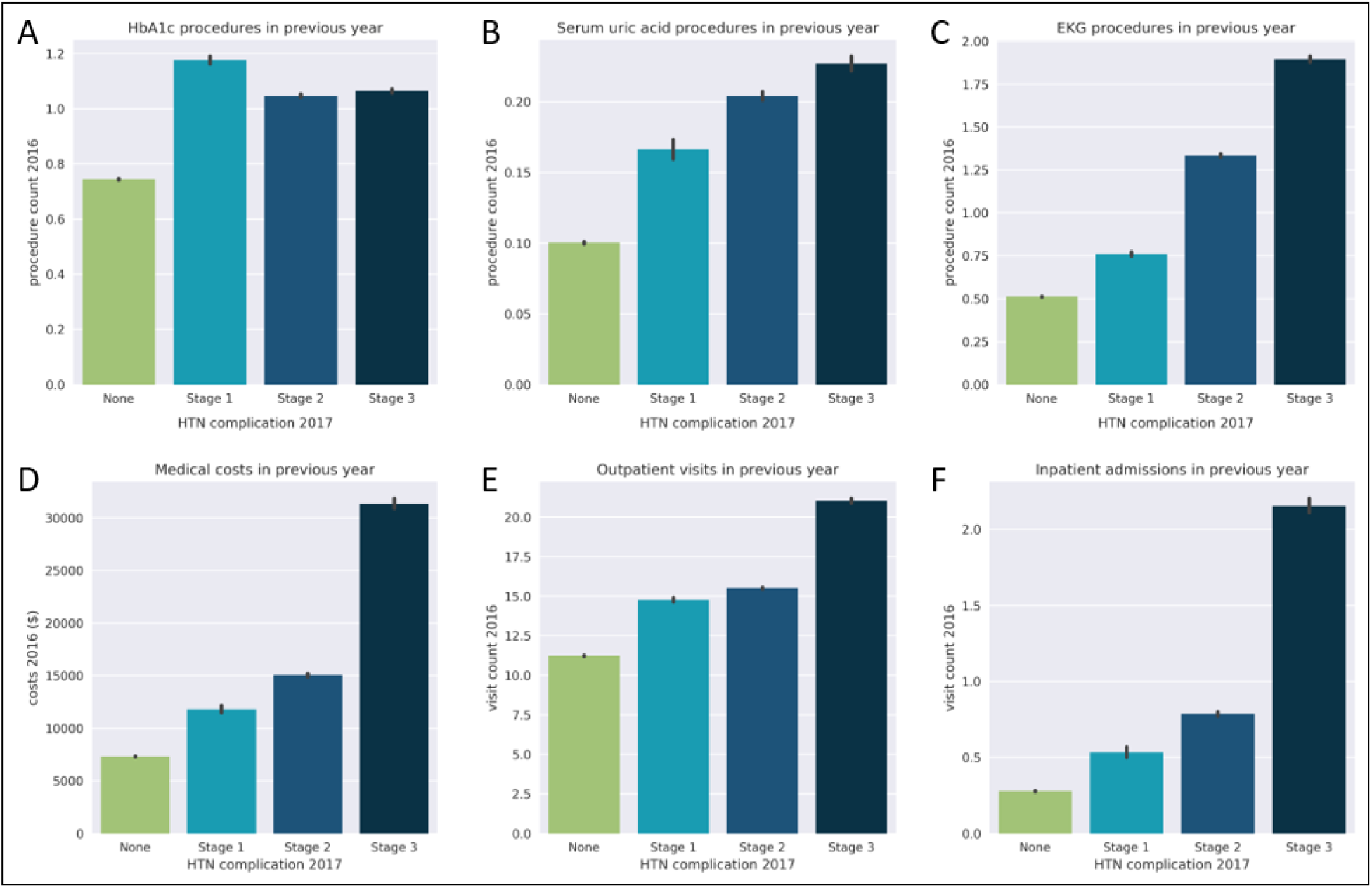
Selected procedure and utilization features. A person is grouped according to his or her most severe complication stage experienced in 2017. (A) Count of procedures for hemoglobin-A1c tests is higher if HTN-related complications experienced in the following year, particularly stage 1 complications. (B-C) Count of procedures for serum uric acid and electrocardiogram procedures increase with increasing complication severity. (D) Total medical spending in previous year increases with complication severity the follow year, particularly for stage 3 complications. (E-F) Number of outpatient visits and inpatient admissions, counted by unique dates of service per member, also increase as HTN-related complications become more serious. Bars represent 95% confidence intervals.

Reducing the overall feature space to two dimensions with principal component analysis for 500 randomly selected cases shows some clustering of no complication cases, with greater spread for stage 2 and stage 3 complications (Supplemental Figure 1).

As some of the complications we are trying to predict, such as chronic kidney disease and dementia, are chronic, we evaluated their presence in our cohort during the intake window and how that may affect model performance. We found that 298,774 (17%) of the cohort had a diagnosis for one of the chronic complication events during the intake year, 216,817 of whom had a complication during the prediction year, accounting for 55% of members of the total cohort that experienced a HTN-related complication during this time. One concern is that the models are benefitting from predicting continued presence of chronic complications; however, not everyone from this group had a complication during the prediction window. Furthermore, models trained on cohorts with and without complication events during the intake window performed less well than when both groups are combined (Supplemental Table S3), so we elected to continue with the latter.

### Prediction performance of ML algorithms

After calculating the features and labels, the data were split randomly into train and test sets with stratified sampling. XGBoost models were fitted and tuned on the training data, and the performance evaluated on the test set, summarized in Supplemental Figure 2. On the independent dataset (Supplemental Figure 3), the model to predict stage 2 complications exhibited the best performance across thresholds (AUROC of 0.89, AUPRC of 0.76). The model to predict stage 3 complications was next best (AUROC of 0.87, AUPRC of 0.58), outperforming the model to predict stage 1 complications (AUROC of 0.81, AUPRC of 0.36) (Figure 5).

**Figure 5.**
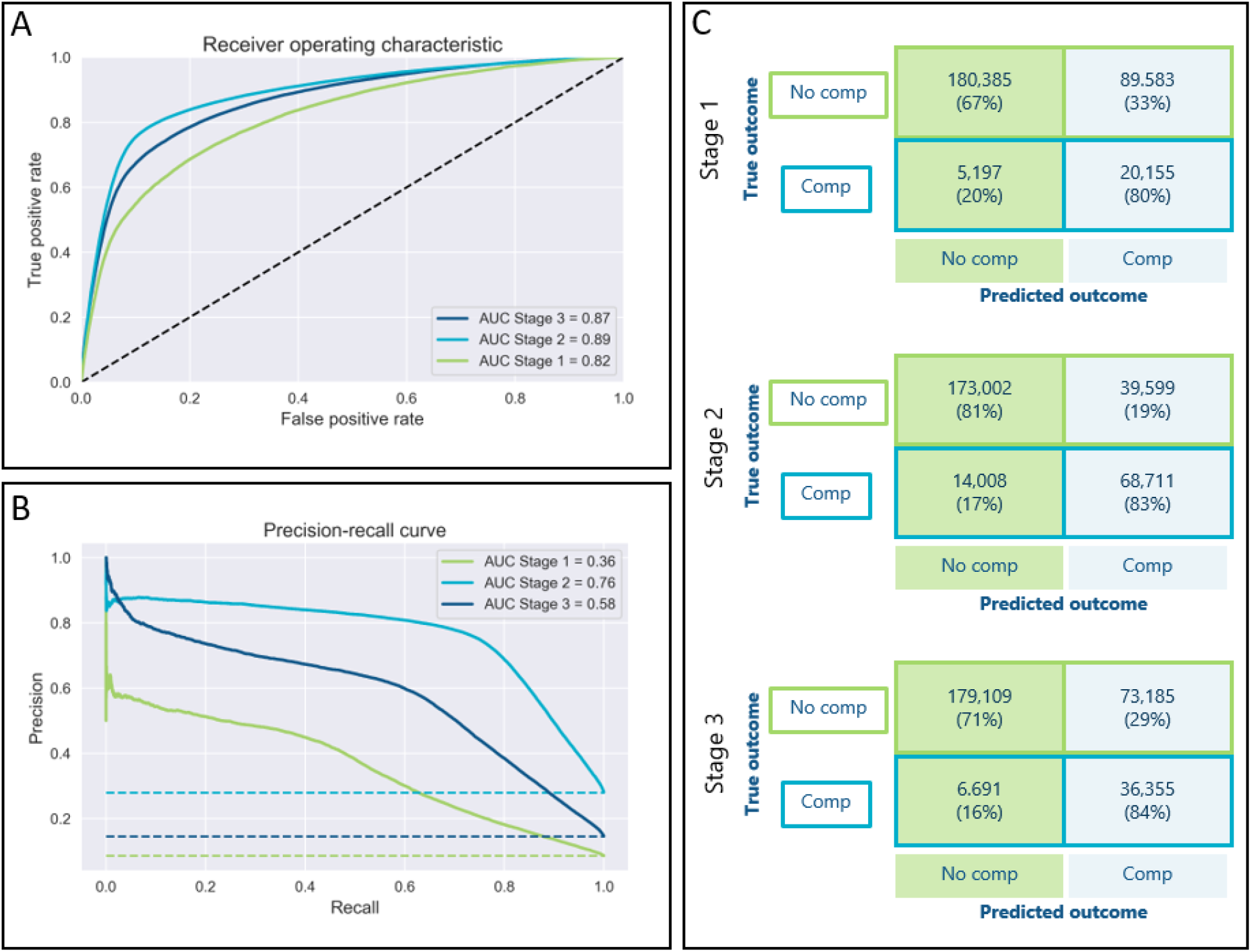
Model performance metrics on independent dataset. (A) Receiving operating characteristic and (B) precision-recall curves for models predicting stage 1, stage 2, and stage 3 complications across thresholds. Dashed lines indicate baseline random guess performance. (C) Confusion matrices based on optimized threshold cutoffs for stage 1, stage 2, and stage 3 models. AUC, area under curve; comp, complication.

At the optimized threshold for specificity and sensitivity as determined by the Youden index, the model for stage 1 complications had a specificity of 66.8%, sensitivity of 79.5%, NPV of 97.2%, PPV of 18.4%, F1 score of 0.30, and NNE of 5.5. Meanwhile, the model for stage 2 complications exhibited a specificity of 81.4%, sensitivity of 93.1%, NPV of 92.5%, PPV of 63.4%, F1 score of 0.72, and NNE of 1.6. Finally, the model for stage 3 complications had a specificity of 71.0%, sensitivity of 84.4%, NPV of 96.4%, PPV of 33.2%, F1 score of 0.48, and NNE of 3.0 (Table 4).

**Table 4.**
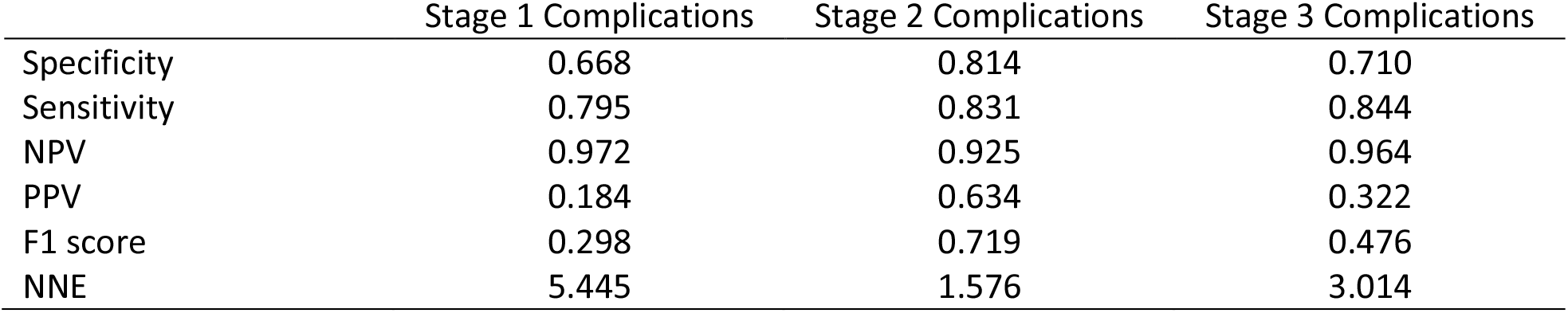
Model performance metrics on independent dataset. Numbers calculated based on threshold cutoffs to optimize specificity and sensitivity on training data. NNE, number needed to evaluate; NPV, negative predictive value; PPV, positive predictive value.

Feature importances were extracted from the XGBoost classifier attributes and the top 20 for each model are shown in Figure 6. The top four predictors across models for all three complication stages were total medical cost, medical cost related to hypertension, outpatient visits, and age, although order varied depending on stage. Other features common to all three models were Charlson Comorbidity Index, count of anti-hypertensive drug classes, count of anti-hypertensive drugs, ED visits, IP visits, EKG count, round of treatment changes, and HbA1c count. Meanwhile, serum uric acid count (stages 1 and 2); valvular disorders and anticoagulants (stages 1 and 3); and sex, statins, beta blockers, breathing difficulties, and thiazide diuretics (stages 2 and 3) were common in two of the three models. Uniquely in the top 20 most important features for predicting stage 1 complications was previous stage 2 and stage 3 complications, edema, diabetes, and atherosclerosis. Unique to stage 2’s top 20 was hyperlipidemia and previous stage 1 complications. For stage 3, dizziness was a top 20 predictor not found for the other two models.

**Figure 6.**
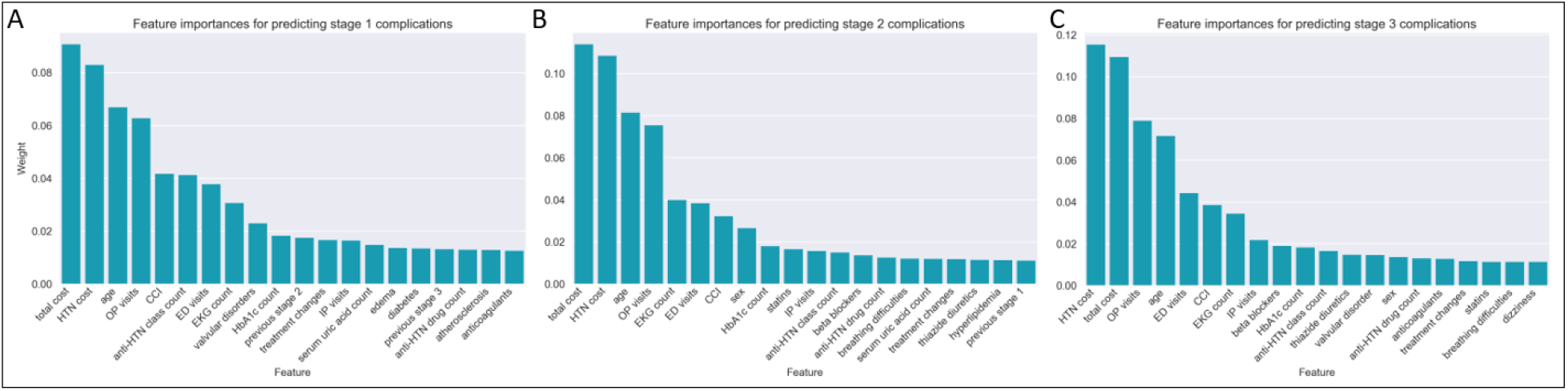
Top 20 most important features for predicting (A) stage 1, (B) stage 2, and (C) stage 3 complications. CCI, Charlson Comorbidity Index; ED, emergency department; EKG, electrocardiogram; HbA1c, hemoglobin A1c; HTN, hypertension; IP, inpatient; OP, outpatient.

## DISCUSSION

There is already a substantial body of literature predicting HTN onset (see [27] for a review), but there remains much value to be added from forecasting HTN-related complications, of which there are more limited studies [9-17]. This would serve the substantial already-diagnosed population, and serve to mitigate the HTN morbidity and mortality burden due to complications. While HTN is associated with major complications such as chronic kidney disease, heart failure, and stroke [28-30], one area that has not been explored to our knowledge is algorithms to predict earlier stage complications. These subtler, sometimes non-specific symptoms, such as microalbuminuria and cognitive dysfunction, may not be recognized as signs of HTN progression like the later stage major complications will be, and yet it is a more effective time to intervene and review treatment and management options.

### Findings

To the above end, we built three separate models to predict different stages of complications that reflect the severity of HTN progression across the target organs of brain, heart, and kidney. On a large commercial population of medical and pharmacy administrative data, we identified prevalence rates of 6%, 17%, and 7%, for stage 1, stage 2, and stage 3 complications, respectively. We trained three XGBoost classifiers using 112 features to predict these different complication stages and found they performed as well or better on an independent dataset than previously published models by AUROC in predicting outcomes most similar to our stage 2 and stage 3 composites [9-13, 15, 17]. The precision or PPV of the models for stages 1 and 3 is low at the threshold chosen to optimize specificity and sensitivity, and could be improved by adjusting the threshold at the sacrifice of other metrics. PPV will always be inherently low for imbalanced datasets, but the NNE of 5.5 and 3.0 for these models is encouraging from an implementation perspective as they reflect a low number of people needed to evaluate to detect one outcome [31].

When looking at the most important features for these models, we found that all five utilization calculated pertaining to cost and visit types (IP, ED, OP) were in the top 20 across models, perhaps because they are good proxies for overall health status. Diagnosis-related features seemed more important in discriminating stage 1 complications (7 features in top 20) than for stage 2 and stage 3 complications (4 features each in the top 20). Conversely, predictors related to medications featured more heavily in the top 20 for stage 2 and stage 3 models (6 and 7 predictors, respectively) than for stage 1 (4 predictors in top 20). A feature to incorporate in the future for all of these models is to look at medication adherence, which has previously been found to increase odds of HTN complications (ischemic heart disease and cerebrovascular disease) [32].

### Care management use cases

Care management for HTN involves educational programs encouraging self-management, including checking blood pressure regularly, understanding the importance of medication adherence even in the absence of symptoms, managing weight and diet, and recognizing signs indicative of HTN-related complications. Should a member become unstable, care managers can coordinate with physicians to schedule appointments, adjust medications, and recommend further lifestyle modifications. Because of the time needed to assess each patient’s unique case, identify and eliminate his or her barriers to care, organize education and resources, and build a trusting relationship, care management assets must be allocated wisely.

Because the number of people with HTN is large and likelihood for HTN-related disease exacerbation is variable, these models can be leveraged to help identify and stratify risk for individuals of a HTN population for care management. People can be rank-sorted by the algorithms’ prediction probability for each complication stage and integrated with other clinical information to assess overall acuity. These predicted complication probabilities could add additional layers for prioritization of outreach in terms of surfacing unique members that may be lower risk by other assessments. The models can also identify patients without previous complication events. Stratification of care management resources could include identifying candidates for remote monitoring programs at home [33, 34], and designating outreach resources ranging from digital strategies to personalized contact with a population health consultant, an engagement specialist, or a clinician. In this way, leveraging these predicative models can help improve the financial and health outcomes impact of care management.

## Limitations

In addition to prevalence issues, part of improving the performance of the stage 1 complication model could entail specialized cohort selection. These models, which were intended to be applied to a hypertension population at large, utilized cohort selection criteria that would have included people at any stage of HTN progression. Since 58% of people that experienced a stage 1 complication also experienced a stage 2 and/or stage 3 complication, the model to predict these early complications is not necessarily fine-tuned to identify progression to these early signs and symptoms. In fact, the presence of stage 1 complications could be continued reporting of these symptoms as people progress to stages 2 and 3. Future iterations of the model will include longer longitudinal data to ensure selection of newly diagnosed HTN patients to improve stage 1 complication detection.

Another limitation of this study is that outside of the diagnoses codes pertaining specifically to hypertension complications (hypertensive heart disease, hypertensive retinopathy, and hypertensive kidney disease), other complication labels selected as part of the composite outcomes cannot necessarily be attributed directly to a person’s hypertension using only administrative data. While using only claims data could be another limitation, even in a clinical setting, determining whether one’s heart attack was due to hypertension or hyperlipidemia or any other combination of factors is not always possible, and why these comorbidities were included as features for prediction. We maintain these models predict risk of events, and their relation to hypertension is important.

## CONCLUSION

Hypertension is a highly prevalent chronic disease, which left unmanaged can progress through increasingly severe complication outcomes, resulting in significant morbidity and mortality. We developed algorithms that could accurately predict different stages of HTN-related complications, grouped by severity level. The ability to predict severity of these outcomes rather than a single composite as most previous models have done allows for more nuanced use cases when these algorithms are utilized in clinical settings.

## Data Availability

The data used for this manuscript are commercially licensed from the CCAE database and not publically available. The independent dataset contains sensitive personal health information and is therefore also not publically available.

## ACKNOWLEDGMENTS

The authors thank Andrew Fairless, Mackenzie DeBoer, and Cindy Meck for helpful discussion.

## SUPPLEMENTARY MATERIAL

**Supplementary Table S1:** HT diagnosis codes and HT medications

| Code type | Code | Description |
| --- | --- | --- |
| ICD-10-CM | I10 | Essential (primary) hypertension |
| ICD-10-CM | I110 | Hypertensive heart disease with heart failure |
| ICD-10-CM | I119 | Hypertensive heart disease without heart failure |
| ICD-10-CM | I120 | Hypertensive chronic kidney disease with stage 5 chronic kidney disease or end stage renal disease |
| ICD-10-CM | I129 | Hypertensive chronic kidney disease with stage 1 through stage 4 chronic kidney disease, or unspecified chronic kidney disease |
| ICD-10-CM | I130 | Hypertensive heart and chronic kidney disease with heart failure and stage 1 through stage 4 chronic kidney disease, or unspecified chronic kidney disease |
| ICD-10-CM | I1310 | Hypertensive heart and chronic kidney disease without heart failure, with stage 1 through stage 4 chronic kidney disease, or unspecified chronic kidney disease |
| ICD-10-CM | I1311 | Hypertensive heart and chronic kidney disease without heart failure, with stage 5 chronic kidney disease, or end stage renal disease |
| ICD-10-CM | I132 | Hypertensive heart and chronic kidney disease with heart failure and with stage 5 chronic kidney disease, or end stage renal disease |
| ICD-10-CM | I150 | Renovascular hypertension |
| ICD-10-CM | I151 | Hypertension secondary to other renal disorders |
| ICD-10-CM | I152 | Hypertension secondary to endocrine disorders |
| ICD-10-CM | I158 | Other secondary hypertension |
| ICD-10-CM | I159 | Secondary hypertension, unspecified |
| ICD-10-CM | I160 | Hypertensive urgency |
| ICD-10-CM | I161 | Hypertensive emergency |
| ICD-10-CM | I169 | Hypertensive crisis, unspecified |
| ATC | C02 | Antihypertensives |
| ATC | C03 | Diuretics |
| ATC | C04 | Peripheral vasodilators |
| ATC | C07 | Beta blocking agents |
| ATC | C08 | Calcium channel blockers |
| ATC | C09 | Agents acting on the renin-angiotensin system |

**Supplementary Table S2:** Complication diagnosis codes

| Stage | Target organ | Broad category | ICD-10-CM code | Description |
| --- | --- | --- | --- | --- |
| Stage 1 | Kidney | Proteinuria | R800 | Isolated proteinuria |
|  |  |  | R801 | Persistent proteinuria, unspecified |
|  |  |  | R802 | Orthostatic proteinuria, unspecified |
|  |  |  | R803 | Bence Jones proteinuria |
|  |  |  | R808 | Other proteinuria |
|  |  |  | R809 | Proteinuria, unspecified |
|  |  | Nephrosclerosis | N269 | Renal sclerosis, unspecified |
|  |  |  | I701 | Atherosclerosis of renal artery |
|  | Heart | LVH | I517 | Cardiomegaly |
|  |  | Atherosclerosis | I70 | Atherosclerosis |
|  | Brain | Retinopathy | H35031 | Hypertensive retinopathy, right eye |
|  |  |  | H35032 | Hypertensive retinopathy, left eye |
|  |  |  | H35033 | Hypertensive retinopathy, bilateral |
|  |  |  | H35039 | Hypertensive retinopathy, unspecified eye |
|  |  | Binswanger lesions | I673 | Progressive vascular leukoencephalopathy |
| Stage 2 | Kidney | Chronic kidney disease | N181 | Chronic kidney disease, stage 1 |
|  |  |  | N182 | Chronic kidney disease, stage 2 (mild) |
|  |  |  | N183 | Chronic kidney disease, stage 3 (moderate) |
|  |  |  | N184 | Chronic kidney disease, stage 4 (severe) |
|  |  |  | N19 | Unspecified kidney failure |
|  |  |  | I129 | Hypertensive chronic kidney disease, stage 1-4 or unspecified |
|  |  |  | I1310 | Hypertensive heart and chronic kidney disease without heart failure with stage 1 through stage 4 chronic kidney disease, or unspecified chronic kidney disease |
|  | Heart | Angina | I200 | Unstable angina |
|  |  |  | I201 | Angina pectoris with documented spasm |
|  |  |  | I208 | Other forms of angina pectoris |
|  |  |  | I209 | Angina pectoris, unspecified |
|  |  | CAD | I2510 | Atherosclerotic heart disease of native coronary artery without angina pectoris |
|  |  |  | I25110 | Atherosclerotic heart disease of native coronary artery with unstable angina pectoris |
|  |  |  | I25111 | Atherosclerotic heart disease of native coronary artery with angina pectoris with documented spasm |
|  |  |  | I25118 | Atherosclerotic heart disease of native coronary artery with other forms of angina pectoris |
|  |  |  | I25119 | Atherosclerotic heart disease of native coronary artery with unspecified angina pectoris |
|  |  |  | I739 | Peripheral vascular disease, unspecified |
|  |  |  | I119 | Hypertensive heart disease without heart failure |
|  |  | Atrial fibrillation | I480 | Paroxysmal atrial fibrillation |
|  |  |  | I481 | Persistent atrial fibrillation |
|  |  |  | I482 | Chronic atrial fibrillation |
|  |  |  | I483 | Typical atrial flutter |
|  |  |  | I484 | Atypical atrial flutter |
|  |  |  | I4891 | Unspecified atrial fibrillation |
|  |  |  | I4892 | Unspecified atrial flutter |
|  |  | Other arrhythmias | I498 | Other specified cardiac arrhythmias |
|  |  |  | I499 | Cardiac arrhythmia, unspecified |
|  | Brain | TIA | G450 | Vertebro-basilar artery syndrome |
|  |  |  | G451 | Carotid artery syndrome (hemispheric) |
|  |  |  | G452 | Multiple and bilateral precerebral artery syndromes |
|  |  |  | G453 | Amaurosis fugax |
|  |  |  | G454 | Transient global amnesia |
|  |  |  | G458 | Other transient cerebral ischemic attacks and related syndromes |
|  |  |  | G459 | Transient cerebral ischemic attack, unspecified |
|  |  | Dementia | F0150 | Vascular dementia without behavioral disturbance |
|  |  |  | F0151 | Vascular dementia with behavioral disturbance |
| Stage 3 | Kidney | ESRD | N185 | Chronic kidney disease, stage 5 |
|  |  |  | N186 | End stage renal disease |
|  |  |  | I120 | Hypertensive chronic kidney disease, stage 5 or ESRD |
|  |  |  | I1311 | Hypertensive heart and chronic kidney disease without heart failure with stage 5 chronic kidney disease or end stage renal disease |
|  |  |  | I132 | Hypertensive heart and chronic kidney disease with heart failure with stage 5 chronic kidney disease or end stage renal disease |
|  | Heart | Ventricular arrhythmias | I4901 | Ventricular fibrillation and flutter |
|  |  |  | I4902 | Ventricular flutter |
|  |  |  | I491 | Atrial premature depolarization |
|  |  |  | I492 | Junctional premature depolarization |
|  |  |  | I493 | Ventricular premature depolarization |
|  |  |  | I4940 | Unspecified premature depolarization |
|  |  |  | I4949 | Other premature depolarization |
|  |  | Myocardial infarction | I2101 | ST elevation (STEMI) myocardial infarction involving left main coronary artery |
|  |  |  | I2102 | ST elevation (STEMI) myocardial infarction involving left anterior descending coronary artery |
|  |  |  | I2109 | ST elevation (STEMI) myocardial infarction involving other coronary artery of anterior wall |
|  |  |  | I2111 | ST elevation (STEMI) myocardial infarction involving right coronary artery |
|  |  |  | I2119 | ST elevation (STEMI) myocardial infarction involving other coronary artery of inferior wall |
|  |  |  | I2121 | ST elevation (STEMI) myocardial infarction involving left circumflex coronary artery |
|  |  |  | I2129 | ST elevation (STEMI) myocardial infarction involving other sites |
|  |  |  | I213 | ST elevation (STEMI) myocardial infarction of unspecified site |
|  |  |  | I214 | Non-ST elevation (NSTEMI) myocardial infarction |
|  |  |  | I219 | Acute myocardial infarction, unspecified |
|  |  |  | I21A1 | Acute myocardial infarction type 2 |
|  |  |  | I21A9 | Other myocardial infarction type |
|  |  |  | I220 | Subsequent ST elevation (STEMI) myocardial infarction of anterior wall |
|  |  |  | I221 | Subsequent ST elevation (STEMI) myocardial infarction of inferior wall |
|  |  |  | I222 | Subsequent non-ST elevation (NSTEMI) myocardial infarction |
|  |  |  | I228 | Subsequent ST elevation (STEMI) myocardial infarction of other sites |
|  |  |  | I229 | Subsequent ST elevation (STEMI) myocardial infarction of unspecified site |
|  |  |  | I230 | Hemopericardium as current complication following acute myocardial infarction |
|  |  |  | I231 | Atrial septal defect as current complication following acute myocardial infarction |
|  |  |  | I232 | Ventricular septal defect as current complication following acute myocardial infarction |
|  |  |  | I233 | Rupture of cardiac wall without hemopericardium as current complication following acute myocardial infarction |
|  |  |  | I234 | Rupture of chordae tendineae as current complication following acute myocardial infarction |
|  |  |  | I235 | Rupture of papillary muscle as current complication following acute myocardial infarction |
|  |  |  | I236 | Thrombosis of atrium, auricular appendage, and ventricle as current complications following acute myocardial infarction |
|  |  |  | I237 | Postinfarction angina |
|  |  |  | I238 | Other current complications following acute myocardial infarction |
|  |  |  | I462 | Cardiac arrest due to underlying cardiac condition |
|  |  |  | I468 | Cardiac arrest due to other underlying condition |
|  |  |  | I469 | Cardiac arrest, cause unspecified |
|  |  | CHF | I420 | Dilated cardiomyopathy |
|  |  |  | I421 | Obstructive hypertrophic cardiomyopathy |
|  |  |  | I422 | Other hypertrophic cardiomyopathy |
|  |  |  | I423 | Endomyocardial (eosinophilic) disease |
|  |  |  | I424 | Endocardial fibroelastosis |
|  |  |  | I425 | Other restrictive cardiomyopathy |
|  |  |  | I426 | Alcoholic cardiomyopathy |
|  |  |  | I427 | Cardiomyopathy due to drug and external agent |
|  |  |  | I428 | Other cardiomyopathies |
|  |  |  | I429 | Cardiomyopathy, unspecified |
|  |  |  | I501 | Left ventricular failure, unspecified |
|  |  |  | I5020 | Unspecified systolic (congestive) heart failure |
|  |  |  | I5021 | Acute systolic (congestive) heart failure |
|  |  |  | I5022 | Chronic systolic (congestive) heart failure |
|  |  |  | I5023 | Acute on chronic systolic (congestive) heart failure |
|  |  |  | I5030 | Unspecified diastolic (congestive) heart failure |
|  |  |  | I5031 | Acute diastolic (congestive) heart failure |
|  |  |  | I5032 | Chronic diastolic (congestive) heart failure |
|  |  |  | I5033 | Acute on chronic diastolic (congestive) heart failure |
|  |  |  | I5040 | Unspecified combined systolic (congestive) and diastolic (congestive) heart failure |
|  |  |  | I5041 | Acute combined systolic (congestive) and diastolic (congestive) heart failure |
|  |  |  | I5042 | Chronic combined systolic (congestive) and diastolic (congestive) heart failure |
|  |  |  | I5043 | Acute on chronic combined systolic (congestive) and diastolic (congestive) heart failure |
|  |  |  | I5081 | Right heart failure |
|  |  |  | I50810 | Right heart failure, unspecified |
|  |  |  | I50811 | Acute right heart failure |
|  |  |  | I50812 | Chronic right heart failure |
|  |  |  | I50813 | Acute on chronic right heart failure |
|  |  |  | I50814 | Right heart failure due to left heart failure |
|  |  |  | I5082 | Biventricular heart failure |
|  |  |  | I5083 | High output heart failure |
|  |  |  | I5084 | End stage heart failure |
|  |  |  | I5089 | Other heart failure |
|  |  |  | I509 | Heart failure, unspecified |
|  |  |  | I130 | Hypertensive heart and chronic kidney disease with heart failure and stage 1 through stage 4 chronic kidney disease, or unspecified chronic kidney disease |
|  |  |  | I110 | Hypertensive heart disease with heart failure |
|  | Brain | Stroke | I60-I66 | Cerebrovascular disease |

**Supplementary Table S3:** Model performance on holdout data for cohorts with and without complications during the intake window Combined refers to cohort including people both with and without complications during the intake window. AUROC, area under the receiver operating characteristic curve; comp, complication; NPV, negative predictive value; PPV, positive predictive value.

|  | Combined stage 1 | With intake stage 1 chronic comp | Without intake stage 1 chronic comp | Combined stage 2 | With intake stage 2 chronic comp | Without intake stage 2 chronic comp | Combined stage 3 | With intake stage 3 chronic comp | Without intake stage 3 chronic comp |
| --- | --- | --- | --- | --- | --- | --- | --- | --- | --- |
| Member count | 1,767,559 | 65,300 | 1,702,259 | 1,767,559 | 240,035 | 1,527,524 | 1,767,559 | 61,534 | 1,706,025 |
| % with comp during prediction window | 5.6 | 30.0 | 4.7 | 17.1 | 72.6 | 8.4 | 7.4 | 66.4 | 5.3 |
| AUROC | 0.791 | 0.670 | 0.757 | 0.879 | 0.749 | 0.759 | 0.850 | 0.757 | 0.787 |
| Specificity | 0.817 | 0.679 | 0.752 | 0.906 | 0.715 | 0.783 | 0.855 | 0.717 | 0.821 |
| Sensitivity | 0.628 | 0.569 | 0.624 | 0.728 | 0.659 | 0.596 | 0.703 | 0.660 | 0.613 |
| NPV | 0.973 | 0.786 | 0.976 | 0.942 | 0.442 | 0.955 | 0.973 | 0.516 | 0.974 |
| PPV | 0.170 | 0.431 | 0.111 | 0.616 | 0.860 | 0.202 | 0.279 | 0.822 | 0.161 |
| F1 score | 0.268 | 0.491 | 0.189 | 0.667 | 0.746 | 0.302 | 0.400 | 0.732 | 0.255 |

## SUPPLEMENTAL FIGURES

**Figure S1.**
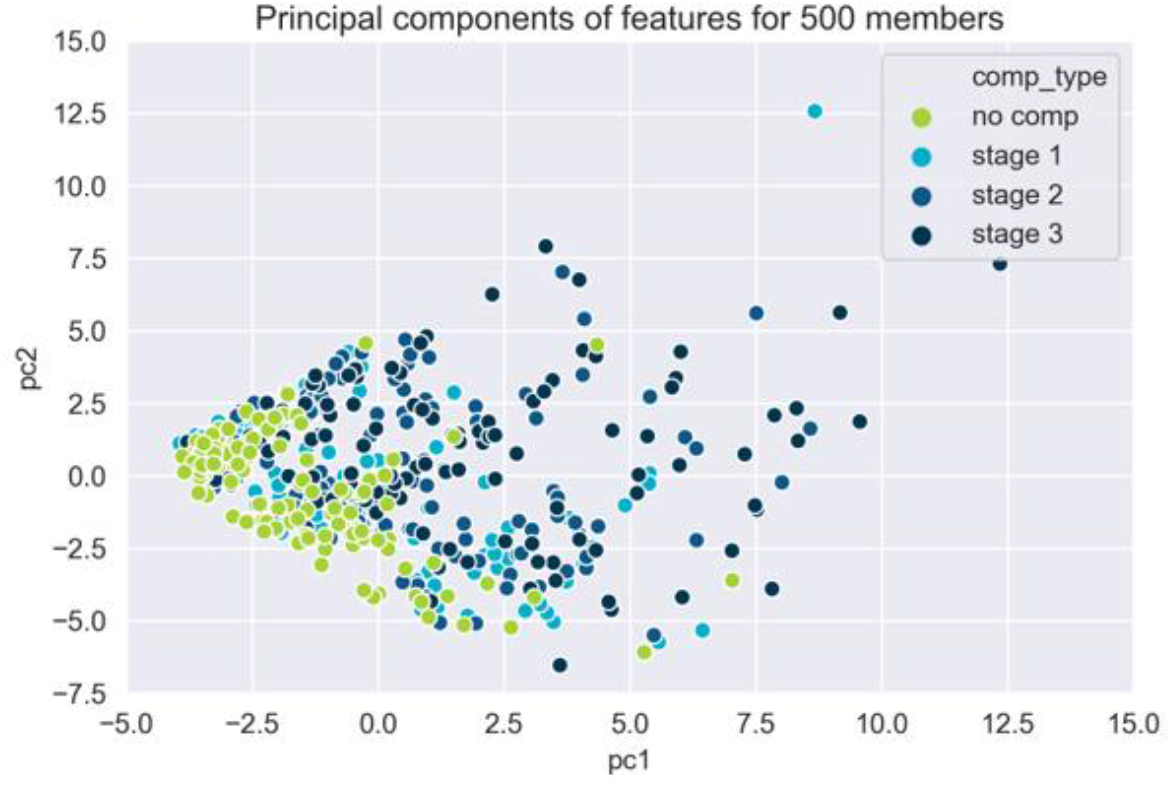
Two dimensional representation of 112 feature inputs with principal component analysis.

**Figure S2.**
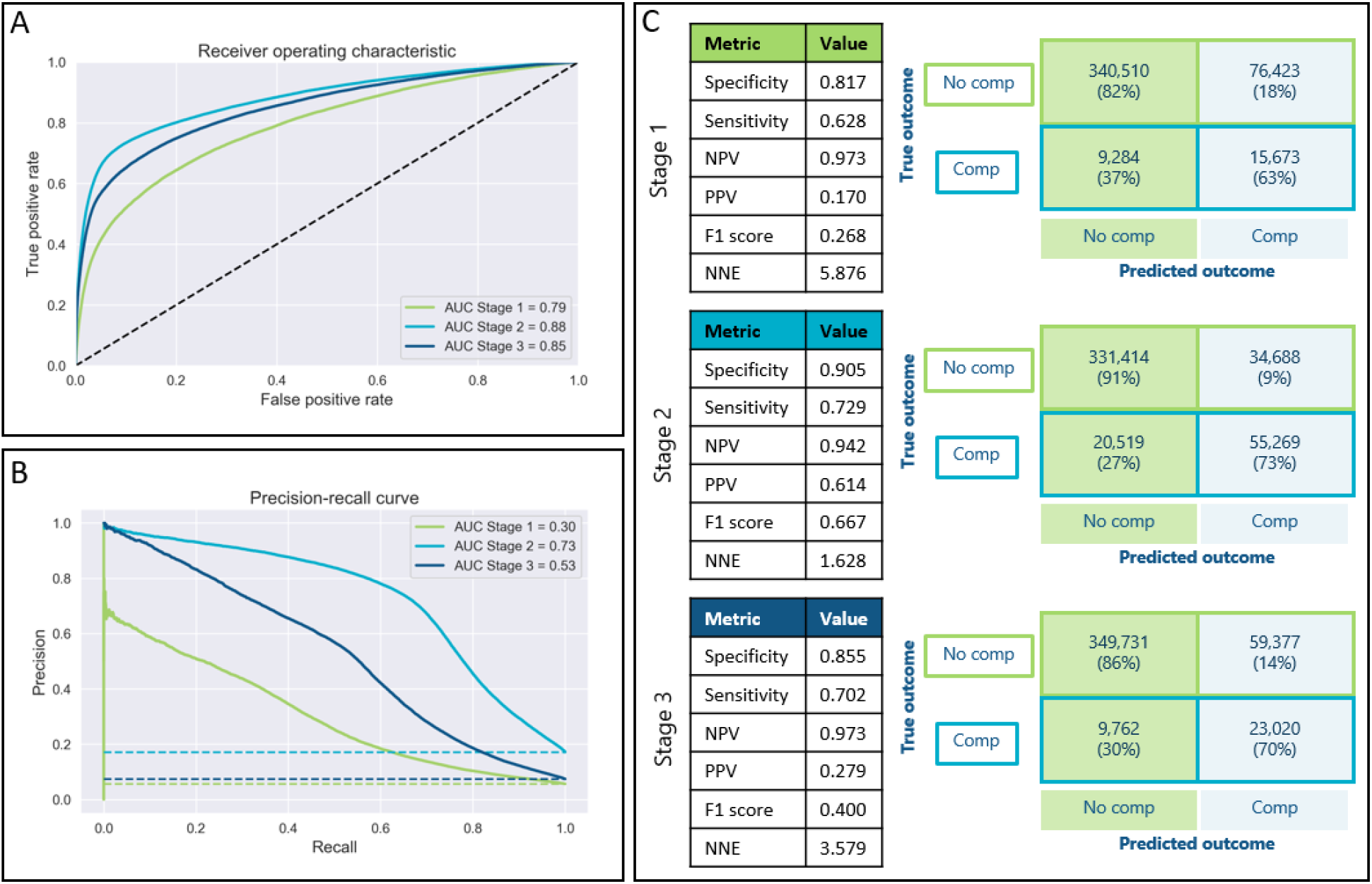
Model performance on Truven holdout dataset. (A) Receiving operating characteristic and (B) precision-recall curves for models predicting stage 1, stage 2, and stage 3 complications across thresholds. Dashed lines indicate baseline performance. (C) Other performance metrics and confusion matrices for stage 1, stage 2, and stage 3 models. AUC, area under curve; comp, complication; NNE, number needed to evaluate; NPV, negative predictive value; PPV, positive predictive value.

**Figure S3.**
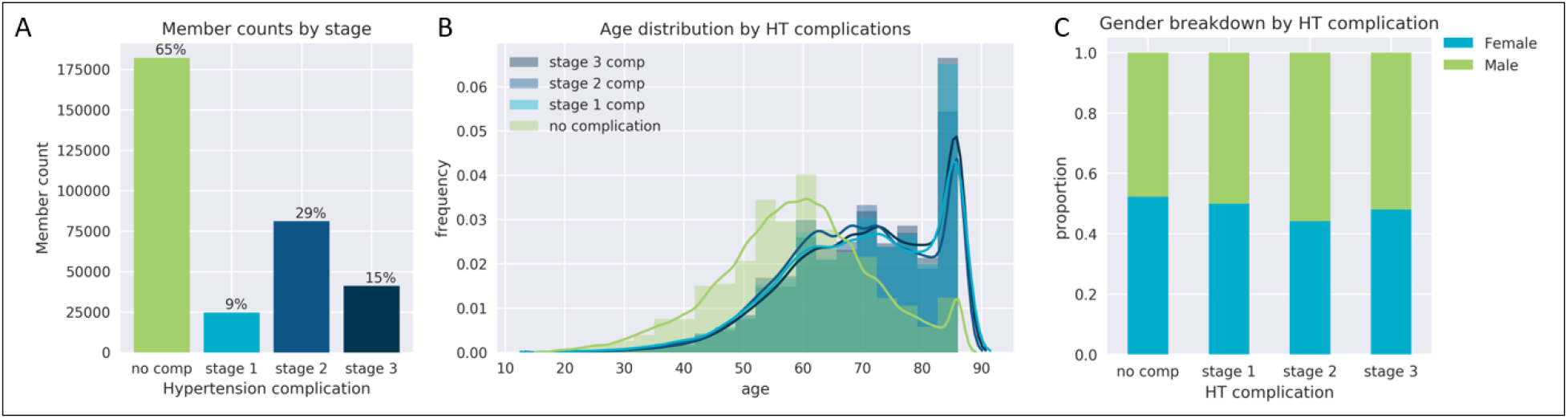
Demographic characteristics of independent dataset population

